# Risk factors for hospital admission related to COVID-19 in inflammatory rheumatic diseases

**DOI:** 10.1101/2020.05.14.20101584

**Authors:** Dalifer Freites, Leticia Leon, Arkaitz Mucientes, Luis Rodriguez-Rodriguez, Judit Font, Alfredo Madrid, Jose Ignacio Colomer, Juan Angel Jover, Benjamin Fernandez-Gutierrez, Lydia Abasolo

## Abstract

**OBJECTIVES:** To describe patients with inflammatory rheumatic diseases (IRD) who had COVID-19; to compare patients who need hospital admission versus those that did not and assess risk factors of hospital admission related to COVID-19.

**METHODS:** We performed a prospective observational study, from 1^st^ March 2020until the 24^th^ of April. All patients being attended at the rheumatology outpatient clinic of a tertiary hospital of Madrid, with medical diagnosis of inflammatory rheumatic disease, and with symptomatic COVID-19 disease were included. Main variable was the hospital admission related to COVID-19. Covariates: sociodemographic, clinical and treatments. We performed a multivariate logistic regression model to assess risk factors of hospital admission.

**RESULTS:** 123 patients with IRD and COVID-19 disease were identified and included. We found

54 patients that need hospital admission, 59.2% were women, with a mean age at hospital admission of 69.7 (15.7) years, and a median lag time from symptoms onset to hospital admission of 5 (3–10) days. The median length of stay was 9 (6–14) days. A total of 12 patients died (22%) during their hospital admission. Factors independently associated with hospital admission were being older (OR 1.08; p = 0.00), and type of diagnosis (OR 3.55; p = 0.01), compared to those who were ambulatory. DMARDs dropped from the model. Male sex, associated comorbidities and glucocorticoids use showed a tendency risk (p<0.2)

**CONCLUSION:** Our results suggests that age, comorbidities and having an autoimmune systemic condition increased the risk of hospital admission, whereas disease modifying agents were not associated with hospital admission.

## INTRODUCTION

Severe acute respiratory syndrome coronavirus 2 (SARS-CoV-2) cause a myriad of clinical signs and symptoms together with analytic typical features. As a whole all characteristics are called COVID-19 disease.^1^

Since the confirmation of the first patient infected with SARS-CoV-2 in Spain in January 2020, the current COVID-19 outbreak has had a great impact, especially in Madrid region, with the highest incidence of COVID-19 cases, with more than 41,304 patients requiring admission at hospitals until the first week of May^2^.

A majority of COVID-19 patients present no symptoms or relatively mild symptomatology. Other smaller but clinically significant subgroup shows progression to a moderate illness. A further subgroup apparently develops a syndrome with autoimmunity and/or autoinflamatory features with critical and/or fatal outcomes.

COVID-19 disease seems to show a higher incidence and severity in patients with risk factors, such as advanced age and some associated comorbidities, mainly hypertension, diabetes, heart disease and previous respiratory diseases^4^. It is not clear if patients with rheumatic diseases have a higher susceptibility to SARS- CoV-2 infection, and when they are infected if they have more severe disease or worse evolution. Based on the clinical information published from previous outbreaks caused by coronaviruses, there is no overwhelming evidence that patients with rheumatic diseases are at an increased risk^3^ although some patients are candidates for a higher number of infections due to their rheumatic disease (predominantly systemic) or their type of treatment for their rheumatic condition^5^. This could be the case of glucocorticoids, that used in a high dose increase the risk of infection. Preliminary experiences on COVID-19 patients show that patients with chronic arthritis treated with biologic or synthetic Disease-modifying antirheumatic drugs (DMARDs) do not seem to be at increased risk of respiratory or life-threatening complications from SARS-CoV-2 compared with the general population^6,7^.

The epidemiological scenario is rapid changing daily and we still have scarce evidence that identify risk factors of poor outcome with COVID-19 specific to inflammatory rheumatic disease, and we have no data on how the hospital admissions of these patients with severe COVID infection have evolved^8^.

The aim of our study is to describe patients with inflammatory rheumatic diseases (IRD) who had COVID-19. Moreover the want to compare patients who need hospital admission versus those that did not, exploring the possible associated risk factors associate to hospital admission related to COVID-19 in IRD patients from a tertiary hospital in Madrid, Spain.

## METHODS

### Setting, Study design, and patients

The setting of this study was a tertiary hospital of the National Health System of the Community of Madrid, Spain, the Hospital Clínico San Carlos (HCSC), covering a catchment area of almost 350,000 people.

We performed a prospective observational study, from the 1^st^ of March 2020, (when our health area had the first hospital admission related to COVID-19) to the 24^th^ of April 2020. All patients being attended at the rheumatology outpatient clinic of our centre during the study period, whose data were recorded in the electronic clinical history of our department (HCR Penelope), were preselected. For the purpose of this study we included patients: **a)** aged > 16 years old; **b)** with medical diagnosis (according to ICD-10) of inflammatory rheumatic disease; and **c)** with symptomatic COVID-19 disease assessed by medical diagnosis or SARS-CoV-2 PCR positive diagnostic test.

Patient data in this project was obtained during routine clinical practice. The study was conducted in accordance with the Declaration of Helsinki and Good Clinical Practices and was approved by the Hospital Clínico San Carlos institutional ethics committee (approval number 20/268-E-BS).

### Variables

The primary outcome was the patient hospital admission with medical diagnosis of COVID-19 and or PCR positive diagnostic test collected from the first of March until the 15^th^ of April compared to those ambulatory patients with symptomatic COVID-19 disease.

The following co-variables were considered: **1)** Sociodemographic baseline characteristics including sex, age and rheumatic disease duration. **2)** Type of inflammatory rheumatic disease diagnosis: including systemic autoimmune conditions (polymyalgia rheumatica; mixed connective tissue disease, systemic sclerosis, sjogren‘s syndrome, vasculitis, Raynaud, polymyositis, polychondritis, sarcoidosis, antiphospholipid syndrome, autoinflammatory syndromes, systemic lupus erythematosus) and chronic inflammatory arthritis (rheumatoid arthritis; inflammatory polyarthritis, juvenile idiopathic arthritis, psoriatic arthritis, axial spondyloarthritis, uvetis, inflammatory bowel disease). 3) Baseline comorbid conditions including: hypertension, dyslipidemia, depression, diabetes mellitus, smoking habit, kidney disease, liver chronic disease, pulmonary disease (Chronic obstructive pulmonary disease and Interstitial Lung Disease), and thyroid disease; heart disease (including valvopathies, arrythmias, myocardiopathies, heart failure or pericarditis); ischemic vascular disease (stroke cardiovascular and peripheral vascular disease), venous thrombosis/lung embolism and cancer disease. **4)** Treatment for inflammatory rheumatic disease: a) glucocorticoids, b) NSAIDs, c) conventional synthetic disease modifying antirheumatic drugs (csDMARDs) including: antimalarials (AM: chloroquine / hydroxychloroquine); sulfasalazine (Ssz); leflunomide (Lef); methotrexate (Mtx); azathioprine or mycophenolato mophetilo (Aza), cyclophosphamide; calcineurin inhibitors (cyclosporine); colchicine; d) targeted synthetic or biologic (ts/bDMARDs), including: d.1) anti-TNF agents (Infliximab, adalimumab, etanercept, certolizumab, golimumab); d.2) Other Biologics: anti-IL6 (tocilizumab, sarilumab); rituximab; anti-IL17 / 23; anti-IL17; abatacept; d.3) Janus kinase inhibitors (tofacitinib, baricitinib).

Treatment had to start at least one month before the beginning of the study, had to continue during the study period until the end of study or medical admission for AM, corticoids, Ssz, NSAIDs, colchicine. Regarding csDMARDs and ts/bDMARDs, treatment had to start at least one month before the beginning of the study, had to continue during at least 21st of March, end of study or medical admission. In the case of Rtx, the last infusion had to be at least on January.

### Data sources

Patient sociodemographic, clinical, laboratory and therapeutic rheumatic data were obtained through the HCR Penelope.

Infected by COVID-19 patients were detected by warning calls to our rheumatologists or nurses or telematic consultation for routine consultations. Other infected patients were obtained through their temporal work disability sick leave forms due to COVID-19. SARS-CoV-2 PCR diagnostic tests were obtained from the microbiology/infectious service of HCSC.

Besides, the Central Services of the Hospital registered all medical admissions of HCSC. All this information was provided from March 1 to April 15.

Of all the hospital admission cases identified in our patients, an exhaustive review of clinical medical records was carried out by the researchers. First of all to identify those medical admissions related to COVID-19 and ruling out those for other reasons. Subsequently, for the COVID-19 cases selected, to collect clinical, laboratory and treatment data during admission until the end of admission (either discharge or death) to describe its evolution. This review was performed until April 24th in order to include follow-up data of those hospital admissions patients with COVID-19.

### Statistical Analysis

Descriptive stats of patient’s characteristics are expressed as mean and standard deviation or median and interquartile rank for continuous variables, while proportions are shown in the case of categorical variables. Statistical tests are performed to compare all patients’ characteristics between those with COVID-19 admissions and those without. Continuous variables have been analyzed using Mann-Whitney or t-student test, and discrete variables have been analyzed using the Chi-square or Fisher exact test. Bivariate logistic regression analyses were done to assess differences between hospital admissions related to COVID-19 risk and covariates. Multivariate logistic regression models (adjusted by age, sex, comorbidity and type of diagnosis) were run to examine the possible risk of sociodemographic, clinical and therapeutic factors on COVID-19 admissions. In the multivariate regression analysis, we have also included comorbidity and DMARDs and all other variables with a p<0.2 obtained in the bivariate analysis. Results were expressed as Odds Ratio (OR) with their respective 95% confidence intervals.

All analyses were performed in Stata v.13 statistical software (Stata Corp., College Station, TX, USA). A two-tailed p value under 0.05 was considered to indicate statistical significance.

## RESULTS

A total of 123 IRD patients with symptomatic COVID-19 disease were included in the study. Table 1 includes a wide sample description.

**Table 1.** Baseline demographic and clinical characteristics among inflammatory rheumatic disease (IRD) patients with COVID-19 (admitted vs no admitted at hospital)

| Variable | IRD Covid-19 patients<br>n =123 | IRD-covid no admitted patients<br>n =69 | IRD-covid admitted patients<br>n =54 | p |
| --- | --- | --- | --- | --- |
| **Gender, women, n (%)** | 86 (69.92) | 54 (78.26) | 32 (59.26) | 0.02 |
| **Age, mean (SD), years** | 59.88 (14.90) | 52.91 (9.58) | 68.78 (15.79) | 0.0001 |
| **Disease evolution time, mean (SD), years** | 10.65 (8.31) | 10.37 (7.99) | 11 (8.77) | 0.8 |
| **PCR test, n (%)** |  |  |  | 0.00 |
| Positive | 58(47) | 17(25) | 41(76) |  |
| Negative | 3(2) | 0 | 3 (5) |  |
| Not performed | 62(51) | 52(75) | 10 (19) |  |
| **Smoking habit (Active versus none)** | 4 (3.25) | 1 (1.45) | 3 (5.56) | 0.31 |
| **Diagnosis, n (%)** |  |  |  | 0.010 |
| Rheumatoid arthritis | 50 (40.65) | 32(46.38) | 18 (33.33) |  |
| Axial spondyloarthritis | 18 (14.63) | 11(15.94) | 7 (12.96) |  |
| Polymyalgia rheumatica | 6 (4.88) | 0 | 6 (11.11) |  |
| Psoriatic arthritis | 6 (4.88) | 3 (4.35) | 3 (5.56) |  |
| Systemic lupus erythematosus | 8 (6.50) | 6 (8.70) | 2 (3.70) |  |
| Mixed connective tissue disease | 6 (4.88) | 2 (2.90) | 4 (7.41) |  |
| Sjogren's syndrome | 9 (7.32) | 5 (7.25) | 4 (7.41) |  |
| Vasculitis | 2 (1.63) | 0 | 2 (3.70) |  |
| Uveitis | 1 (0.81) | 1 (1.45) | 0 |  |
| Systemic sclerosis | 1 (0.81) | 0 | 1 (1.85) |  |
| Inflammatory polyarthritis | 8 (6.50) | 6 (8.70) | 2 (3.20) |  |
| Polychondritis | 1(0.81) | 0 | 1 (1.85) |  |
| Polymyositis | 1(0.81) | 0 | 1 (1.85) |  |
| Raynaud | 3 (2.44) | 0 | 3 (5.56) |  |
| Others* | 3 (2.44) | 3 (4.35) | 0 |  |
| **Comorbidities, n (%)** |  |  |  |  |
| Hypertension | 40 (32.52) | 14 (20.29) | 26 (48.15) | 0.002 |
| Dyslipidemia | 27 (21.95) | 12(17.35) | 15 (27.38) | 0.19 |
| Depression | 9 (7.32) | 8 (11.59) | 1 (1.85) | 0.039 |
| Diabetes Mellitus | 17 (13.82) | 4 (5.80) | 13 (24.07) | 0.007 |
| Heart disease | 15(12.20) | 5 (7.25) | 10 (18.52) | 0.09 |
| Vascular disease | 8 (6.50) | 2 (2.90) | 6 (11.11) | 0.13 |
| Liver disease | 7 (5.69) | 3 (4.35) | 4 (7.41) | 0.69 |
| Kidney disease | 6 (4.88) | 0 | 6 (11.11) | 0.006 |
| Lung disease (ILD/COPD) | 19 (15.45) | 6 (8.70) | 13 (24.07) | 0.02 |
| Cancer | 5 (4.07) | 1 (1.45) | 4 (7.41) | 0.16 |
| Venous Thrombosis/lung embolism | 3 (2.44) | 0 | 3 (5.56) | 0.08 |
| Thyroid disease | 17(13.8) | 12 (17.39) | 5 (9.26) | 0.29 |
| **NSAIDs, n (%)** | 30 (24.39) | 22 (31.88) | 8 (14.81) | 0.03 |
| **Glucocorticoid use, n (%)** | 61 (49.59) | 29 (42.03) | 32 (59.26) | 0.07 |
| **csDMARDs, n (%):** |  |  |  |  |
| Mtx-Lef-Aza | 68 (55.28) | 40 (57.97) | 28 (51.85) | 0.49 |
| Slz | 9 (7.32) | 5 (7.25) | 4 (7.41) | 0.97 |
| AM | 27 (21.95) | 18 (26.09) | 9 (16.67) | 0.21 |
| **bDMARDs, n (%)** | 26 (21.14) | 19 (27.54) | 7 (12.96) | 0.04 |
| Anti-TNF | 17 (13.82) | 15 (21.74) | 2 (3.70) | 0.004 |
| **Non Anti-TNF** | 9 (7.32) | 4 (5.80) | 5(9.26) | 0.4 |
| Abata | 1 (0.81) | 1(1.45) | 0 | 0.99 |
| Tozi | 2 (1.63) | 1(1.45) | 1(1.85) | 0.99 |
| Beli | 1 (0.81) | 1(1.45) | 0 | 0.00 |
| Rtx | 5 (4.07) | 1(1.45) | 4 (7.41) | 0.16 |
| **JAKi , n (%)** | 1 (0.89) | 0 | 1 (2) | 0.43 |

**Abbreviations:** IRD: inflammatory rheumatic disease. SLE: systemic lupus erythematosus; RA: rheumatoid arthritis IA: inflammatory polyarthritis PSA: psoriatic arthritis; SPA: spondyloarthritis; PMR: polymyalgia rheumatic; SSc: systemic sclerosis; CTMD: mixed connective tissue disease; Sjo: Sjögren's syndrome. Others\*: Inflammatory bowel disease, antiphospholipid syndrome, juvenile idiopathic arthritis, autoinflammatory syndromes; sarcoidosis. \*\*Heart disease: arrhythmias, valvulopathies, cardiomyopathies, heart failure. Ischemic vascular disease: stroke, cardiovascular and peripheral vascular disease. ; COPD: chronic obstructive pulmonary disease; ILD: interstitial lung disease. ts/bDMARDs target synthetic/biologic disease-modifying anti rheumatic drug; Anti-TNF: Tumor necrosis factor-alpha inhibitor; JAKi: JAK inhibitors; csDMARD: Conventional synthetic disease-modifying anti rheumatic drug. Other Bios: anti-IL6 (tocilizumab, sarilumab); rituximab (Rtx); anti-IL17 / 23; anti-IL17; JAKi: JAK inhibitors.

Most of the patients were women with a mean age of 59.88 (14.9) years, and disease evolution mean time of 10.65 (8.31) years. The main diagnosis was rheumatoid arthritis (40.65% of the patients), being followed by axial spondyloarthritis, with the 14.63% of the sample. An elevated number of patients had at least one basal comorbid condition being hypertension, dyslipidemia, and lung disease the most prevalent ones. Most patients were taking csDMARDs at the beginning of study (71.54%). Half of the patients were taking glucocorticoids (49.59%) and a quarter of the patients were taking NSAIDs (24.39%). A 21.14% were taking biologic DMARDs (bDMARDs). The most frequently bDMARDs used was Adalimumab (6.50%), followed by Rituximab (4.07%). Only one patient was taking a Janus kinase inhibitors (Tofacitinib). Interestingly, 14.63% of the patients on bDMARDs were in combined therapy with a sDMARD.

We found a total of 54 patients from our cohort that needed hospital admission because COVID-19. Of those patients, 51 were evaluated in our Emergency department (ED) at HCSC, and finally 49 were admitted at our hospital. Other two patients were referred to IFEMA support hospital due to lack of capacity in our hospital at that time. In addition, 3 patients were admitted to other regional hospitals (Figure 1).

**Figure 1.**
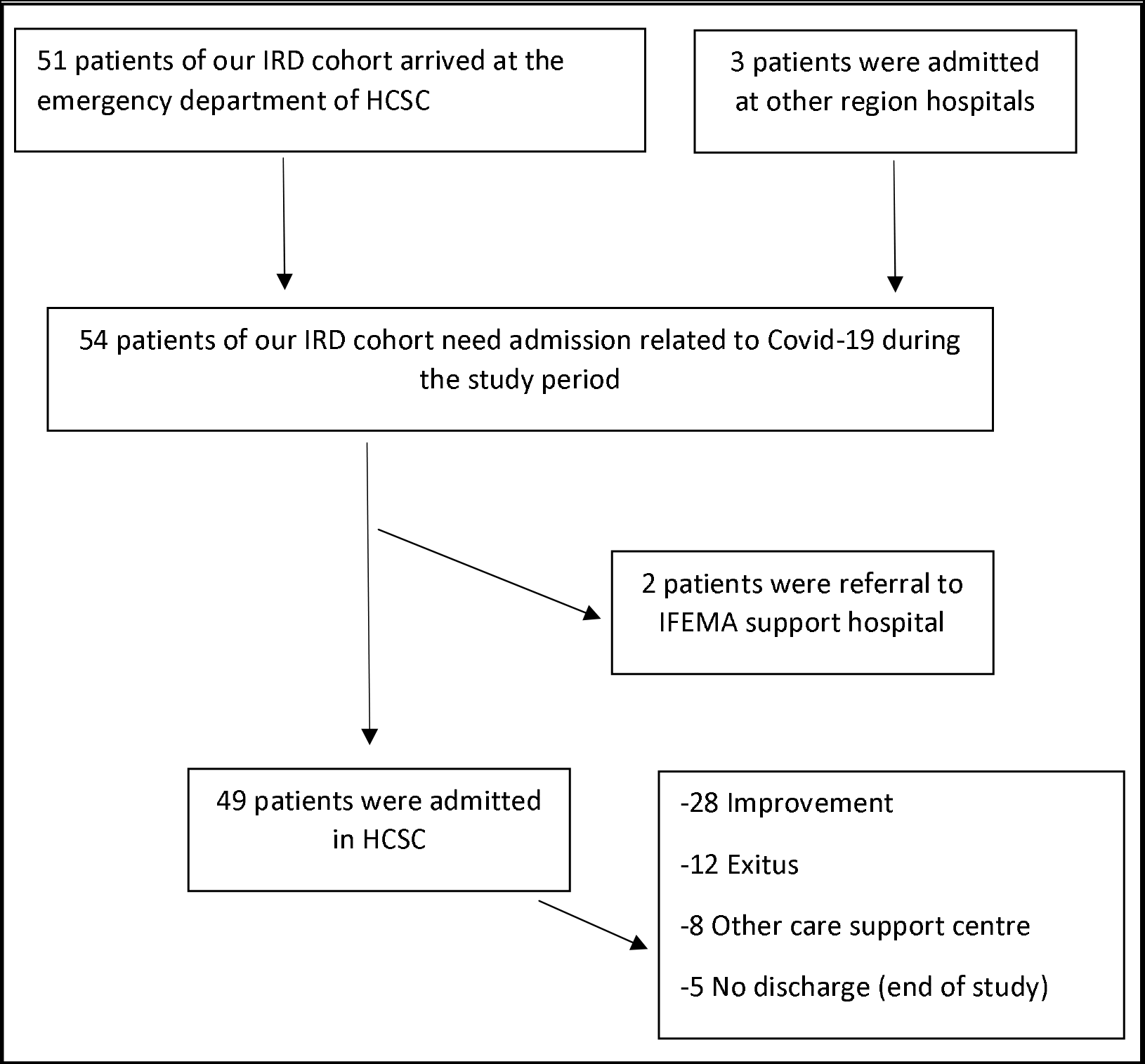
Flow chart for COVID-19 patient hospital admission

Of the patients admitted to our hospital, 59.2% of them were in women, with a mean age at admission of 69.7 (15.7) years, and a median lag time from symptoms onset to admission of 5(3–10) days. The median length of stay was 9 (6–14) days (Table 2).

**Table 2.** Hospital admission related to Covid-19 among IRD patients.

| Variable | Value |
| --- | --- |
| **Number of admissions recorded in ED, n\*** | 51 |
| **Number of admissions communicated, n\*** | 3 |
| **Number of admissions at hospital, n** | 49 |
| **Women, n (%)** | 31 (58.49) |
| **Age at admission, median (IQR)** | 72.5 (53-82) |
| **Lag time from symptoms onset to admission, days, median (IQR)** | 5 (3-10) |
| **Length of stay, days, median (IQR)** | 9 (6-14) |
| **Discharge reason, n (%)** |  |
| Improvement, home isolation | 29 (53.70) |
| Other care center (medicalized hotel/IFEMA hospital) | 8 (14.82) |
| Exitus | 12 (22.22) |
| End of study (no discharge) | 5 (9.26) |
| **Main baseline comorbidities, n (%)** |  |
| Hypertension | 26 (48.15) |
| Dyslipidemia | 15 (27.78) |
| Diabetes mellitus | 13 (24.07) |
| Cancer | 4 (7.41) |
| Cardiovascular disease | 6 (11.11) |
| Lung diseases | 13 (24.07) |
| Chronic liver disease | 4 (7.41) |
| Chronic renal disease | 6 (11.11) |
| **Pneumonia at admission, n (%)** | 47 (87) |
| **Systemic autoimmune conditions, n (%)** | 24 (44,4) |
| **Admitted by Intensive Care Unit during hospital admission** |  |
| No | 52 (96.29) |
| Yes | 2 (3.71) |
| **Laboratory data at admission, median (IQR)** |  |
| Hemoglobin g/dl | 12.9 (12.4-13.8) |
| D-dimer, ng/ml | 727 (487-1091) |
| Neutrophil count, $\times 10^9/L$ | 4,500 (3,500-5,700) |
| Lymphocyte count, $\times 10^9/L$ | 700 (500-1,200) |
| CRP mg/dl | 9.19 (2.9-14.6) |
| LDH, u/l | 618 (489-919) |
| Platelet count $\times 10^9/L$ | 199,000(158,000-267,000) |
| Creatinine, mg/dl | 0.86 (0.76-1.28) |
| Ferritine, ng/ml | 319 (151-885) |
| **Covid-19 related treatments during admission, n (%)** |  |
| Azitromicine | 17 (34) |
| Other antibiotic | 29 (58) |
| Glucocorticoids | 26 (52) |
| Lopinavir / Ritonavir | 18 (6) |
| Remdesivir | 0 |
| Darunavir/cobicistat | 4 (8) |
| Tocilizumab | 3 (6) |
| Interferon inhalation | 4 (8) |
| HCQ | 43 (86) |
| Immunoglobulin | 0 |

**Abbreviations:** ED: Emergency department; SD: standard deviation; IQR: Interquartile range; HCQ: Hidroxychloroquine. \*51 were evaluated in our Emergency department (ED) at HCSC, finally 49 were admitted at our hospital. Other two patients were referred to IFEMA support hospital due to lack of capacity in our hospital at that time. In addition, 3 patients were admitted to other regional hospitals (a third contacted our rheumatology service to report the disease)

At the time of admission, the median Hemoglobin was 12.9 (p25 12.4-p75 13.8), and the median total lymphocyte count was 700 (p25 500-p75 1,200). The median for D-dimer was 727 (p25 487–75 1,091). Our patients have been given different antibiotics (mainly Azithromycin,

Levofloxacine and third generation cephalosporines).

Most patients were treated with Hydroxychloroquine during the admission (86%). About half of the patients received corticosteroids (52%). 18 patients were treated with Lopinavir / Ritonavir and 3 patients received anti–IL-6R antibody tocilizumab. Other characteristics are shown in Table 2. Of those 49, five went to another care center (medicalized hotel / IFEMA support hospital) when their condition improves.

A total of 20 patients (44%) developed relevant complications during admission, the most frequent being myocarditis, thrombosis or kidney failure. Only two patients were admitted by Intensive Care Unit during hospital admission. The first was a 55-year-old man with a MCTD and associated comorbidities that developed an acute respiratory insufficiency and bilateral pneumonia. He was treated with antibiotic therapy, Lopinavir/ritonavir, hidroxychroloquine and β- Interferon. After 40 days he was extubated and he is he continues to recover. Other patient was a 29-year-old woman with SLE, treated with Mtx, Rtx, hydroxychloroquine and glucocorticoids, that days after their COVID-19 diagnosis (PCR +), developed an erythematous rash and generalized urticaria, requiring hospitalization in intensive care unit due to general clinical and laboratory worsening, with D-dimer elevated. She was treated with Methylprednisolone, heparin and cephalosporine. One week after she improved and was discharge completely recovered.

Of those admitted in our hospital, five went to another care center (medicalized hotel / IFEMA support hospital) when their condition improves. Other 29 patients (53.7%) were discharged and went home to continue with home isolation after improvement. At the end of study, five patients were still admitted (9.26%).A total of 12 patients died (22%) during the admission (6 males and 6 females), with a median age of 81 (76.5–87) years, being the main diagnosis of patients RA (6), following by SPA (2), PMR (2), vasculitis (1) and Sjogren‘s syndrome (1).

The bivariate analysis is shown in table 3. Older age, systemic autoinmune conditions compared to chronic inflammatory arthritis, hypertension, diabetes mellitus, lung disease increased and heart disease the risk of hospital admission with statistical signification. Whereas being women, the use of NSAIDs and TNF-targeted bDMARD compared to non-TNF-targeted bDMARD had less risk with statistical signification. Glucocorticoids had a tendency of higher risk and AM had a tendency of lower risk of hospital admission.

**Table 3.**
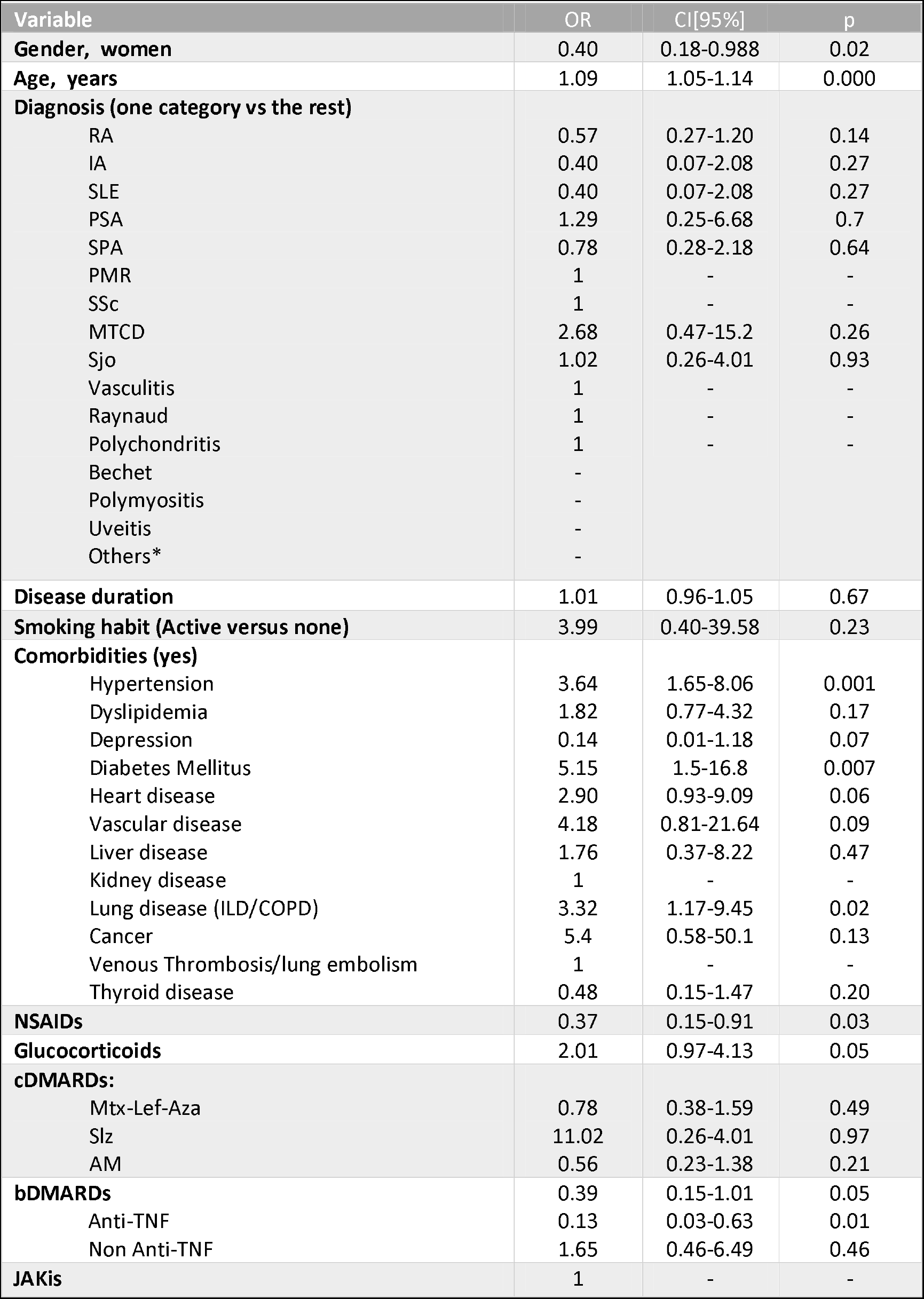
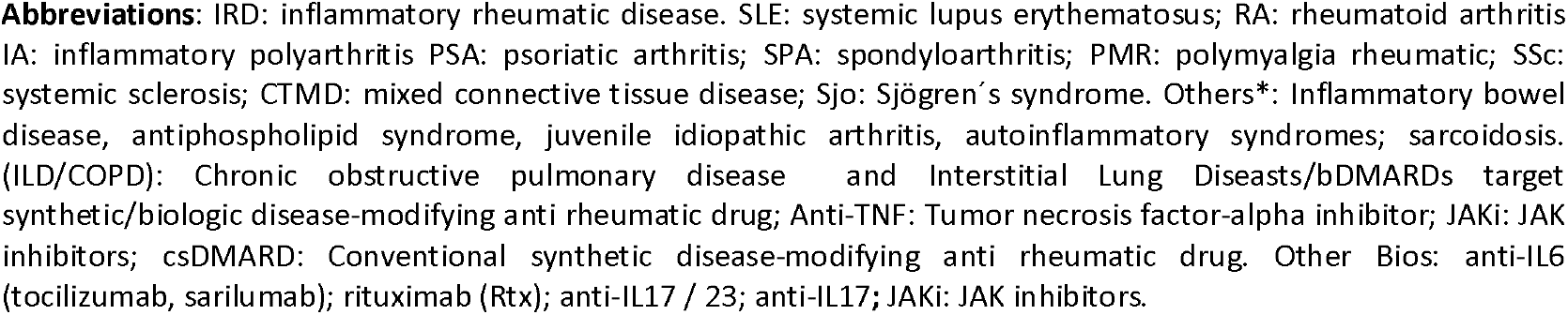
Odd ratio of medical admission related to COVID-19 in IRD patients. Bivariate analysis.

| Variable | OR | CI[95%] | p |
| --- | --- | --- | --- |
| **Gender, women** | 0.40 | 0.18-0.988 | 0.02 |
| **Age, years** | 1.09 | 1.05-1.14 | 0.000 |
| **Diagnosis (one category vs the rest)** |  |  |  |
| RA | 0.57 | 0.27-1.20 | 0.14 |
| IA | 0.40 | 0.07-2.08 | 0.27 |
| SLE | 0.40 | 0.07-2.08 | 0.27 |
| PSA | 1.29 | 0.25-6.68 | 0.7 |
| SPA | 0.78 | 0.28-2.18 | 0.64 |
| PMR | 1 | - | - |
| SSc | 1 | - | - |
| MTCD | 2.68 | 0.47-15.2 | 0.26 |
| Sjo | 1.02 | 0.26-4.01 | 0.93 |
| Vasculitis | 1 | - | - |
| Raynaud | 1 | - | - |
| Polychondritis | 1 | - | - |
| Bechet | - |  |  |
| Polymyositis | - |  |  |
| Uveitis | - |  |  |
| Others* | - |  |  |
| **Disease duration** | 1.01 | 0.96-1.05 | 0.67 |
| **Smoking habit (Active versus none)** | 3.99 | 0.40-39.58 | 0.23 |
| **Comorbidities (yes)** |  |  |  |
| Hypertension | 3.64 | 1.65-8.06 | 0.001 |
| Dyslipidemia | 1.82 | 0.77-4.32 | 0.17 |
| Depression | 0.14 | 0.01-1.18 | 0.07 |
| Diabetes Mellitus | 5.15 | 1.5-16.8 | 0.007 |
| Heart disease | 2.90 | 0.93-9.09 | 0.06 |
| Vascular disease | 4.18 | 0.81-21.64 | 0.09 |
| Liver disease | 1.76 | 0.37-8.22 | 0.47 |
| Kidney disease | 1 | - | - |
| Lung disease (ILD/COPD) | 3.32 | 1.17-9.45 | 0.02 |
| Cancer | 5.4 | 0.58-50.1 | 0.13 |
| Venous Thrombosis/lung embolism | 1 | - | - |
| Thyroid disease | 0.48 | 0.15-1.47 | 0.20 |
| **NSAIDs** | 0.37 | 0.15-0.91 | 0.03 |
| **Glucocorticoids** | 2.01 | 0.97-4.13 | 0.05 |
| **cDMARDs:** |  |  |  |
| Mtx-Lef-Aza | 0.78 | 0.38-1.59 | 0.49 |
| Slz | 11.02 | 0.26-4.01 | 0.97 |
| AM | 0.56 | 0.23-1.38 | 0.21 |
| **bDMARDs** | 0.39 | 0.15-1.01 | 0.05 |
| Anti-TNF | 0.13 | 0.03-0.63 | 0.01 |
| Non Anti-TNF | 1.65 | 0.46-6.49 | 0.46 |
| **JAKis** | 1 | - | - |

**Abbreviations:** IRD: inflammatory rheumatic disease. SLE: systemic lupus erythematosus; RA: rheumatoid arthritis IA: inflammatory polyarthritis PSA: psoriatic arthritis; SPA: spondyloarthritis; PMR: polymyalgia rheumatic; SSc: systemic sclerosis; CTMD: mixed connective tissue disease; Sjo: Sjögren's syndrome. Others\*: Inflammatory bowel disease, antiphospholipid syndrome, juvenile idiopathic arthritis, autoinflammatory syndromes; sarcoidosis. (ILD/COPD): Chronic obstructive pulmonary disease and Interstitial Lung Diseases/bDMARDs target synthetic/biologic disease-modifying anti rheumatic drug; Anti-TNF: Tumor necrosis factor-alpha inhibitor; JAKi: JAK inhibitors; csDMARD: Conventional synthetic disease-modifying anti rheumatic drug. Other Bios: anti-IL6 (tocilizumab, sarilumab); rituximab (Rtx); anti-IL17 / 23; anti-IL17; JAKi: JAK inhibitors.

In the multivariate analysis after adjusting by gender, age, comorbidities and type of diagnosis (Table 4), we did not find statistical difference between type of DMARDs. In fact AM (p = 0.6) other cDMARDs (p = 0.7), NSAID (p = 0.5), TNF (p = 0.17) and Anti-TNF (p = 0.3) dropped from the model. Patients with older age (p = 0.00), and systemic autoimmune conditions (p = 0.01) had more probability of hospital admissions regardless other factors. Glucocorticoids (p = 0.15), the presence of comorbidity related to COVID-19 (defined as having at least one of the follows: diabetes mellitus, pulmonary disease, ischemic vascular disease, hypertension, venous thrombosis/lung embolism, lung disease and or liver disease) (p = 0.2) and being women (p = 0.1) did not reached statistical signification, although both showed a trend.

**Table 4.** Multivariate adjusted analysis of risk factors for admission in IRD patients.

| Variable | OR | CI[95%] | p |
| --- | --- | --- | --- |
| Gender, women | 0.45 | 0.15-1.29 | 0.1 |
| Age , years | 1.08 | 1.04-1.13 | 0.00 |
| Diagnosis (Systemic autoimmune conditions vs chronic inflammatory arthritis) | 3.55 | 1.30-9.67 | 0. 01 |
| COVID Comorbidities (yes) | 1.82 | 0.69-4.80 | 0.2 |
| Glucocorticoids | 1.97 | 0.77-5.01 | 0.15 |

## DISCUSSION

This study aims to shed some light on rheumatologists about their concerns regarding their patients. Our data showed that, in a real-world setting, there is a 44% of inflammatory rheumatic patients with COVID-19 that required hospital admission, mainly elderly patients, with more comorbidities and systemic autoimmune condition. This study provides additional evidence that patients treated with disease modifying agents are not at a higher risk of hospital admission related to COVID-19.

Our admitted patients had a median age near the 70, being more than fifteen years older than patients who were not admitted. Patients who did not recovery and finally died had a median age over the 80. This is in line with general population data, where over 95% of these deaths occurred in those older than 60 years and more than 50% of all deaths were people aged 80 years or older^7^.

Regarding the probability of admission, despite the fact that rheumatic diseases have a higher prevalence in women, we have not found a higher risk of admission in this group. The type of diagnosis seems to play an important role in the probability of hospital admission, having those patients with systemic autoimmune conditions the highest risk.

Our admitted patients had the same laboratory results as those of other published studies previously described^23^. Biomarkers predictive of poor outcomes have included lymphopenia and elevations in C-reactive protein (CRP), interleukin (IL)-6, and D-dimer, among others^10–12^.

Some comorbidities play an important role in the risk of hospital admission. It has already been seen that COVID-19 patients with any comorbidity yielded poorer clinical outcomes than those without and a greater number of comorbidities also correlated with poorer clinical outcomes^13^. The importance of diabetes as comorbidity in COVID-19 has already been seen, finding that the history of diabetes are independent risk factors for morbidity and mortality in COVID-19^14,15^. Diabetes has been previously associated with of admissions to the intensive care unit (ICU) due to COVID-19 in recent series^16,17^. It also seems to increase mortality^6^.

Our results on glucocorticoids must be taken with caution because we analyzed if they were using or not regardless the dosage and they may refer to dose-dependent patients.In a recent study including almost 40,000 patients diagnosed with polymyalgia rheumatica or giant cell arteritis, it was found high absolute risks of all types of infection and a marked stepwise increase in risk with higher oral glucocorticoid doses^18^. Previously, a study has demonstrated a dose-related relationship between prednisone treatment and the risk of pneumonia in rheumatoid arthritis patients in the community, however no increased risk was found with anti-TNF therapy or methotrexate^19,20^.

Our results found that chronic exposition to DMARDs- do not seem to present a more risk of hospital admission risk related to COVID-19, neither biologic nor synthetic DMARDs.

Regarding the treatments they received during admission, it has been very varied in patients since the disease has been a challenge for specialists, who have been outlining different combinations of drugs with not too much published evidence.

The use of interleukin 6 (IL-6) blockers seems to be very promising for the management of the massive cytokine storm associated to the development of the typical lung damage and the consequent ARDS occurring in the most aggressive patterns of SARS-CoV infection^21^.

IL-6 levels have been reported to be elevated in admitted patients related to COVID-19, and also it has been reported that the anti–IL-6R antibody tocilizumab has been beneficial in COVID-19 patients^22^. Treatment also may be successful in early stages of cytokine release syndrome (CRS). If they can effectively block the signal transduction pathway of IL-6, thus tocilizumab and sarilumab are likely to emerge as effective drugs for patients with moderate to severe COVID-19^23^.

The results of our study should be interpreted considering limitations. First, we have to take into account the observational nature of the study and that our patients were treated at a single center. This, associated with the fact that data was recorded during routine consultations, that it is an environment with heavy workload, makes easier the possibility of incomplete and not recoverable information. Another limitation is the difficulty to identify patients. Of the patients who did not require admission, a third contacted the rheumatology service to report the disease, and the other cases were detected through the discharge parts by COVID-19 of their primary care physician. For sure, elderly or housewives who have not contacted us are missing, and may have patient selection biases between those admitted and not admitted, however this has been solved adjusting for confounders in multivariate analysis.

PCR test should be required for main outcome definition. However, in all admissions during this period, almost 20% of them did not have PCR performed due to a lack of available tests or extreme health care overload. Nevertheless, all cases included were carefully reviewed being clinically compatible and managed as COVID-19.

Finally, we have not been able to study if ethnic differences have a role in the severity COVID-19, but doctors believe that there are a large number of infected people from some groups that for economic or cultural reasons tend to share a home with more people.

However, studies including IRD patients with COVID-19 are still limited, but the study of Haberman et al show similar results^8^. Nevertheles, both studies have small sample size and more evidence is desirable in order to clarify prognostic factors about these patients.

These data can help rheumatologists to treat and advise their patients on this new challenge, as well as to know which of the factors may be associated with an increased risk of hospital admission in patients with rheumatic inflammatory diseases.

## Data Availability

Data are available upon request

## Acknowledgements

The authors would also like to thank Ana M Perez for their help in the data collection. A special thank you to all rheumatologist and nurses colleagues who contributed in the care of the patients in an innovative and so involved way.

## Contributors

BF, LL, JAJ, LRR and LA contributed to the conception and design of the study. DF, JF, AM, JIC and LL were involved in data collection. LA and LL performed the data analysis and interpretation of data. All authors contributed to drafting and/or revising the manuscript.

## Ethics approval

The study was approved by the Hospital Clínico San Carlos institutional ethics committee (approval number 20/268-E-BS). This study was conducted according to the principles of the Declaration of Helsinki.

## Patient and Public Involvement

Patients or the public were not involved in the design, or conduct, or reporting, or dissemination plans of our research.

